# Medical student residency preferences and motivational factors: a longitudinal, single-institution perspective

**DOI:** 10.1101/2022.01.29.22270073

**Authors:** Feria A. Ladha, Anthony M. Pettinato, Adam E. Perrin

## Abstract

**Background:** A high proportion of medical school graduates pursue specialties different from those declared at matriculation. While these choices influence the career paths, satisfaction, and potential regret students will experience, they also impact the supply and demand ratio of the shorthanded physician workforce across many specialties. In this study, we investigate how the choice of medical specialty and the factors motivating those choices change between the beginning and end of medical school training.

**Methods:** A questionnaire was administered annually from 2017 to 2020 to a cohort of medical students at the University of Connecticut to determine longitudinal preferences regarding residency choice, motivational factors influencing residency choice, future career path, and demographic information.

**Results:** The questionnaire respondent totals were as follows: n=76 (Year 1), n=54 (Year 2), n=31 (Year 3), and n=65 (Year 4). Amongst newly matriculated students, 25.0% were interested in primary care, which increased ∼1.4-fold to 35.4% in the final year of medical school. In contrast, 38.2% of matriculated students expressed interest in surgical specialties, which decreased ∼2.5-fold to 15.4% in the final year. Specialty choices in the final year that exhibited the largest absolute change from matriculation were orthopedic surgery (−9.9%), family medicine (+8.1%), radiology (+7.9%), general surgery (−7.2%), and anesthesiology (+6.2%). Newly matriculated students interested in primary care demonstrated no differences in their ranking of motivational factors compared to students interested in surgery, but many of these factors significantly deviated between the two career paths in the final year. Specifically, students interested in surgical specialties were more motivated by the rewards of salary and prestige compared to primary care students, who more highly ranked match confidence and family/location factors.

**Conclusions:** We identified how residency choices change from the beginning to the end of medical school, how certain motivational factors change with time, how these results diverge between primary care and surgery specialty choice, and propose a new theory based on risk-reward balance regarding residency choice. Our study promotes awareness of student preferences and may help guide school curricula in developing more student-tailored training approaches. This could foster positive long-term changes regarding career satisfaction and the physician workforce.

## Background

For medical students, deciding on a residency specialty that will guide careers and impact personal lives is a complicated and multifactorial process, made even more difficult by the complex and stressful nature of attempting to switch residency specialties post hoc (1). These decisions have broad implications regarding healthcare and biomedical research across the globe, namely an imbalance between physician supply and demand in primary care (2–4), surgery (5–7), research (8–10), and clinical subspecialties (11–13), a disparity that dates back over a century and will continue to worsen for the foreseeable future (14–16). These disparities can have a significant impact on healthcare outcomes. For example, a greater supply of primary care physicians per capita is associated with improved cardiovascular health, lower mortality, increased lifespan, and a reduction in low birth-weight rates (17–19). While the physician workforce landscape is shaped by numerous variables that can differ in importance and oversight across municipalities, states/provinces, and countries, principal persistent factors that contribute are the personal desires and interests of the physician trainees themselves, which may or may not align with the needs of the healthcare system (20).

Investigating which specialties medical students prefer and why has been of long-standing interest to medical schools and healthcare administrations, as a better understanding of these preferences could provide insights that enable improvements to education curricula that better foster the wants and needs of future physicians and the healthcare system. Previous studies have demonstrated that the residency preferences of matriculating medical students change by the time a final residency choice is made at the end of medical school (21,22). Notably, students have demonstrated a persistent tendency to categorically switch preference between primary care and surgical specialties (23–26). While informative studies have observed specific predictive factors influencing residency choice, such as demographics, interest, lifestyle, finances, and prestige, the single time point nature of these studies limits our understanding of how these factors may change over time (23–25,27–30), and results can be further confounded due to recall bias from methods that require retrospective assessment by respondents. Moreover, alterations to the medical curriculum itself have been shown to impact residency choice (31–34), making it imperative to obtain accurate and current data regarding student preferences that can be used to facilitate optimal curriculum changes.

In this longitudinal study, we track the residency specialty preferences and motivational factors for a cohort of U.S. medical students throughout their training at the University of Connecticut School of Medicine. The aim of this study is to identify if and how the residency preferences of newly matriculated medical students change compared to the residency specialties chosen in their last year of medical school. Concurrently, this study also aims to investigate the factors influencing residency choice, with a focus on how these factors may or may not change with time and between specialty categories, such as between primary care and surgical specialties. The results from this study will add to the growing understanding of medical student career preferences. This may help to inform decisions regarding medical education, such as more personalized training plans that foster career satisfaction and guidance towards in-demand specialties.

## Methods

### Subjects and Questionnaire

This study was reviewed by the Institutional Review Board (IRB) at UConn Health and qualified for exempt status (IRB number 18-062-3). A voluntary, self-administered questionnaire (**Table 1**) was developed to longitudinally assess medical students’ residency specialty preferences, as well as the factors motivating those preferences, throughout their medical school training towards conferral of their medical degree (M.D.). A survey was chosen as the data collection instrument in order to measure attitudes/beliefs, as these qualities are personal/internal and thus not directly observable. The questionnaire was administered annually from 2017 to 2020 to the same student cohort from the University of Connecticut School of Medicine (UConn SOM Class of 2021), a 4-year M.D.-granting medical school in the U.S., starting upon matriculation (Year 1; n=102 matriculants) and concluding during submission of residency applications using the Electronic Residency Application Service (ERAS) (Year 4; n=100 matches). The anonymous questionnaire consisted of respondents self-reporting their preferred (Years 1-3) or chosen (Year 4) residency specialty, their likely career path following residency (e.g. subspecialty training, research, etc.), and ranking the various motivational factors (e.g. specialty lifestyle, prestige, etc.) on these stated choices, in addition to demographics information (e.g. age, marital status, etc.). To meet recent guidelines for survey-based research, this questionnaire format (**Table 1**) is similar to what has been utilized in previous studies (22,23,36), was reviewed by two faculty members unaffiliated with the study, and a survey trial with interview-based feedback was performed using a small number of respondents outside the cohort of interest to ensure content, face, and response process validity as well as interrater reliability (35,37).

**Table 1.** Survey questionnaire. Simplified questionnaire presenting the questions and response options for the students to complete.

| Questions | Response Options |
| --- | --- |
| What residency are you most likely to pursue following medical school? (choose one) | List of residency specialties (see Table 3) |
| What career path(s) are you most likely to pursue following residency? (choose any/all that apply) | -Fellowship/Subspecialty Training<br>-Research<br>-Industry |
| Rank the following six factors in terms of importance to you in deciding your specialty/career choice (1=least important and 6=most important). | -Family and/or location<br>-Interest in the field itself (e.g. underlying science, day-to-day duties, target patient population, subsequent training opportunities, etc.)<br>-Prestige<br>-Lifestyle (e.g. hours)<br>-Financial incentive (e.g. salary)<br>-Residency match confidence (e.g. ease of matching) |
| Degree program (choose one) | MD - MD/PhD - MD/MBA - MD/MPH - other |
| Current year (choose one) | MS1 - MS2 - MS3 - MS4 |
| Age | (fill in) |
| Are you married? | Yes - No |
| Do you have children? | Yes - No |

### Procedure

For Years 1-3, the questionnaire was administered in-person at curriculum sessions in which a majority of students would be present, such as at the end of lecture (Years 1-2) or prior to an orientation session (Year 3) and students were given 20 minutes to complete the survey. The surveyors (F.A.L. and A.M.P.) announced the goals and voluntary/anonymous nature of the study to the students present, distributed paper copies of the questionnaire, and collected the completed questionnaires with respect to respondent anonymity. Due to the COVID-19 pandemic, the Year 4 questionnaire was distributed using the class email listserv and voluntary, anonymous responses were submitted electronically. No incentive was offered for completing the survey.

### Data Analysis

The data obtained in this study, including the internally distributed results from the National Resident Matching Program (NRMP) for the UConn SOM (n=100), were electronically cataloged and analyzed. Data in this study excluded oral and maxillofacial surgery and preliminary surgery Match outcomes, as well as longitudinal results from students in dual degree programs (e.g. M.D./Ph.D., M.D./M.B.A, etc), as these represent atypical training timelines and/or career paths. For the “primary care” specialty categorization, this encompassed internal medicine, pediatrics, and family medicine, as defined by the American Academy of Family Physicians (AAFP). For the “surgery” specialty categorization, this encompassed general surgery, orthopedic surgery, neurological surgery, vascular surgery, plastic surgery, otolaryngology, and urology, as defined by the American College of Surgeons (ACS). Obstetrics and gynecology (OB/GYN) was excluded from either classification due to its hybrid nature and evolving landscape (38–41).

With the potential for small sample sizes due to specific residency choice, incomplete questionnaires, and/or survey distribution logistics (i.e. cohort availability), a 90 percent confidence level (α = 0.1) was chosen (42,43). For analysis, the percentage of students who chose each residency specialty were calculated relative to the total respondents for each year the questionnaire was completed. To assess correlation between Year 4 survey results and Match outcomes, a Pearson correlation coefficient (*r*) and its associated *P*-value were calculated using GraphPad Prism. The importance of the six factors motivating the chosen specialties, which were numerically recorded on an ordinal scale from least important (1) to most important (6), were compiled by both medical school year and specialty choice. The median scale values and interquartile ranges were calculated and plotted in GraphPad Prism. Statistical comparisons were performed using nonparametric Mann-Whitney U tests in GraphPad Prism, and the resulting *P*-values were considered statistically significant if *P* ≤ 0.1.

## Results

The total number of cohort respondents by year were n=76 for Year 1, n=54 for Year 2, n=31 for Year 3, and n=64 for Year 4, as summarized in **Table 2**. Due to the lower response rate in Years 2 and 3, we directed our focus and analyses on Years 1 and 4. The age range for Year 1 respondents was 21-33 years-old and 25-34 years-old for Year 4 respondents. This was accompanied by an increase in the percentage of married respondents (6.6% in Year 1 and 15.6% in Year 4) and respondents with children (0.0% in Year 1 and 3.1% in Year 4).

**Table 2.** Respondent demographics. This table includes the number of student volunteers who completed the questionnaire in each of the four years it was administered, as well as the demographic data they provided.

| Medical School Year | Number of Respondents | Age Range of Respondents | Married Respondents | Respondents with Children |
| --- | --- | --- | --- | --- |
| Year 1 | 76 | 21-33 | 5 (6.6%) | 0 (0.0%) |
| Year 2 | 54 | 22-30 | 3 (5.6%) | 0 (0.0%) |
| Year 3 | 31 | 23-30 | 1 (3.2%) | 0 (0.0%) |
| Year 4 | 65 | 25-34 | 10 (15.4%) | 2 (3.1%) |

With regard to residency specialty choices (**Table 3**), the top six choices for the entering students in Year 1 were internal medicine (14.5%), emergency medicine (14.5%), orthopedic surgery (14.5%), general surgery (11.8%), pediatrics (7.9%), and OB/GYN (7.9%). For Year 4, the top six residency specialty choices were internal medicine (15.4%), emergency medicine (12.3%), family medicine (10.8%), pediatrics (9.2%), OB/GYN (9.2%), and radiology (9.2%). Of note, the specialty choices that exhibited the greatest absolute change from Year 1 to Year 4 were orthopedic surgery (−9.9%), family medicine (+8.1%), radiology (+7.9%), general surgery (−7.2%), and anesthesiology (+6.2%). This correlated well with the final NRMP Match outcomes for this cohort of UConn SOM students (*r* = 0.91, *P* < 0.001), as the greatest absolute changes from Year 1 to the Match were orthopedic surgery (−12.5%), anesthesiology (+8.0%), general surgery (−7.8%), radiology (+6.7%), and family medicine (+5.4%; tied with psychiatry). Of note, the least popular specialties—those that garnered no interest in Year 1 and no Match outcomes—were child neurology, combined internal medicine and pediatrics (med-peds), pathology, and physical medicine and rehabilitation. Finally, the majority of students reported a desire to pursue fellowship/subspecialty training after residency in both Year 1 (51.3%) and Year 4 (66.2%), whereas 0% of Year 1 students were interested in research compared to 13.8% of students in Year 4 (**Table 4**). Industry was the least likely post-residency path for respondents, with only 2.6% of Year 1 students and 4.6% of Year 4 students expressing interest.

**Table 3.** Residency specialty results. This table represents the number of students reporting their preferred residency specialty (Years 1-3), chosen residency specialty (Year 4), and Match outcome for this cohort. The percentage for each response/result by year is also provided.

| Residency | Year 1<br>Response<br>(n=76) | Year 2<br>Response<br>(n=54) | Year 3<br>Response<br>(n=31) | Year 4<br>Response<br>(n=65) | NRMP Match<br>Outcome<br>(n=100) |
| --- | --- | --- | --- | --- | --- |
| Anesthesiology | 0 (0.0%) | 0 (0.0%) | 0 (0.0%) | 4 (6.2%) | 8 (8.0%) |
| Child Neurology | 0 (0.0%) | 1 (1.9%) | 0 (0.0%) | 0 (0.0%) | 0 (0.0%) |
| Dermatology | 3 (4.0%) | 0 (0.0%) | 0 (0.0%) | 2 (3.1%) | 4 (4.0%) |
| Emergency<br>Medicine | 11 (14.5%) | 10 (18.5%) | 3 (9.7%) | 8 (12.3%) | 12 (12.0%) |
| Family Medicine | 2 (2.6%) | 4 (7.4%) | 2 (6.5%) | 7 (10.8%) | 8 (8.0%) |
| General Surgery | 9 (11.8%) | 5 (9.3%) | 4 (12.9%) | 3 (4.6%) | 4 (4.0%) |
| Internal Medicine | 11 (14.5%) | 11 (20.4%) | 5 (16.1%) | 10 (15.4%) | 15 (15.0%) |
| Med-Peds | 0 (0.0%) | 0 (0.0%) | 0 (0.0%) | 0 (0.0%) | 0 (0.0%) |
| Neurosurgery | 3 (4.0%) | 1 (1.9%) | 1 (3.2%) | 1 (1.5%) | 0 (0.0%) |
| Neurology | 3 (4.0%) | 4 (7.4%) | 1 (3.2%) | 4 (6.2%) | 4 (4.0%) |
| OB/GYN | 6 (7.9%) | 2 (3.7%) | 0 (0.0%) | 6 (9.2%) | 8 (8.0%) |
| Ophthalmology | 0 (0.0%) | 2 (3.7%) | 0 (0.0%) | 0 (0.0%) | 2 (2.0%) |
| Orthopedic<br>Surgery | 11 (14.5%) | 2 (3.7%) | 1 (3.2%) | 3 (4.6%) | 2 (2.0%) |
| Otolaryngology | 3 (4.0%) | 4 (7.4%) | 2 (6.5%) | 2 (3.1%) | 4 (4.0%) |
| Pathology | 0 (0.0%) | 0 (0.0%) | 0 (0.0%) | 1 (1.5%) | 0 (0.0%) |
| Pediatrics | 6 (7.9%) | 4 (7.4%) | 8 (25.8%) | 6 (9.2%) | 9 (9.0%) |
| Physical Med &<br>Rehab | 0 (0.0%) | 0 (0.0%) | 0 (0.0%) | 0 (0.0%) | 0 (0.0%) |
| Plastic Surgery | 1 (1.3%) | 0 (0.0%) | 1 (3.2%) | 0 (0.0%) | 1 (1.0%) |
| Psychiatry | 2 (2.6%) | 2 (3.7%) | 3 (9.7%) | 1 (1.5%) | 8 (8.0%) |
| Radiation<br>Oncology | 2 (2.6%) | 1 (1.9%) | 0 (0.0%) | 0 (0.0%) | 0 (0.0%) |
| Radiology | 1 (1.3%) | 1 (1.9%) | 0 (0.0%) | 6 (9.2%) | 8 (8.0%) |
| Urology | 0 (0.0%) | 0 (0.0%) | 0 (0.0%) | 1 (1.5%) | 3 (3.0%) |
| Vascular Surgery | 2 (2.6%) | 0 (0.0%) | 0 (0.0%) | 0 (0.0%) | 0 (0.0%) |

**Table 4.** Career paths following completion of residency. Students were asked to choose which career path they were most likely to follow after completion of residency. Students were given the option to choose multiple pathways if they wished.

| Medical School Year | Fellowship/<br>Subspecialty Training | Research | Industry |
| --- | --- | --- | --- |
| Year 1 | 39 (51.3%) | 0 (0.0%) | 2 (2.6%) |
| Year 2 | 32 (59.3%) | 8 (14.8%) | 2 (3.7%) |
| Year 3 | 21 (67.7%) | 5 (16.1%) | 3 (9.7%) |
| Year 4 | 43 (66.2%) | 9 (13.8%) | 3 (4.6%) |

To determine which factors may motivate a student’s preferred/chosen residency specialty, we asked respondents to rank six factors—family/location, field interest, prestige, lifestyle, financial incentive, and match confidence—from least important to most important in motivating their reported residency specialty choice. Using a linear scale (1=least important to 6=most important) to numerically compare these responses, we identified no significant leading motivational factor in Year 1, as the median ranking was relatively similar across the six assessed factors (**Figure 1**). However, when compared to Year 4, significant changes were observed. Interest in the field itself (e.g. underlying science, day-to-day duties, target patient population, subsequent training opportunities, etc.) and field lifestyle (e.g. hours) significantly increased in importance in Year 4, whereas field prestige, financial incentive (e.g. salary), and match confidence significantly decreased in importance. The importance of family and/or location requirements did not significantly change from Year 1 to Year 4.

**Figure 1.**
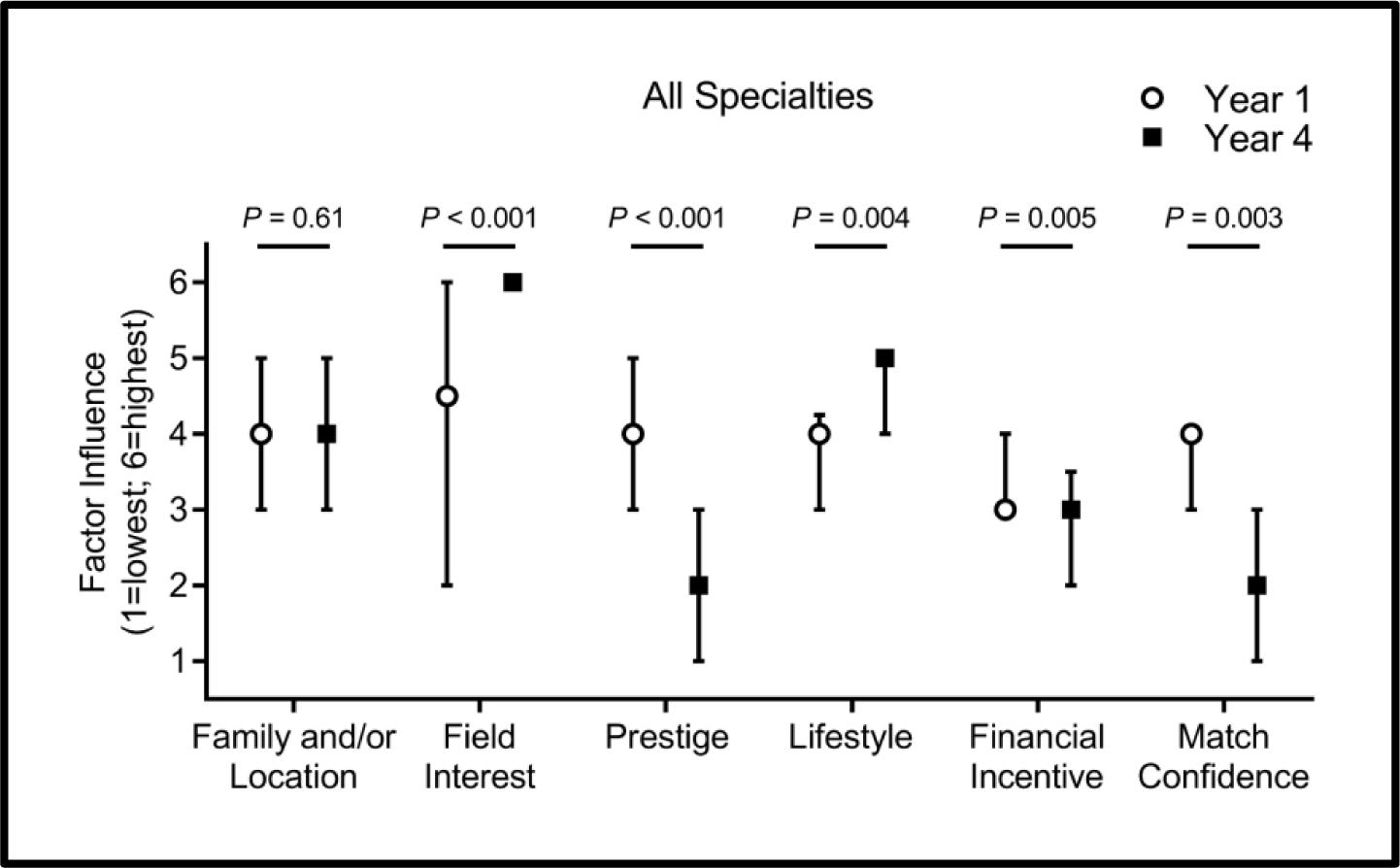
Factors motivating residency choice. Median and interquartile range for the six motivational factors influencing choice of residency specialties from least important (1) to most important (6), comparing the student cohort in Year 1 (white circle) versus Year 4 (black square). *P*-values were calculated using Mann-Whitney U tests.

As we observed disparities and changes between specialty choices within and between Years 1 and 4, we next set out to compare interest amongst broad specialty categorizations, specifically primary care versus surgery, as change between these specialty categories during medical school has been shown to occur across studies from multiple decades (23–25,33,34). In Year 1, 25.0% of students were interested in primary care specialties (internal medicine, pediatrics, or family medicine), which increased ∼1.4-fold to 35.4% in Year 4 and ∼1.3-fold to 32.0% in the final Match outcome (**Figure 2A**). In contrast, 38.2% of Year 1 students expressed interest in surgical specialties (orthopedic surgery, general surgery, neurological surgery, vascular surgery, plastic surgery, otolaryngology, and urology), which decreased ∼2.5-fold to 15.4% in Year 4 and ∼2.7-fold to 14.0% in the final Match outcome (**Figure 2B**).

**Figure 2.**
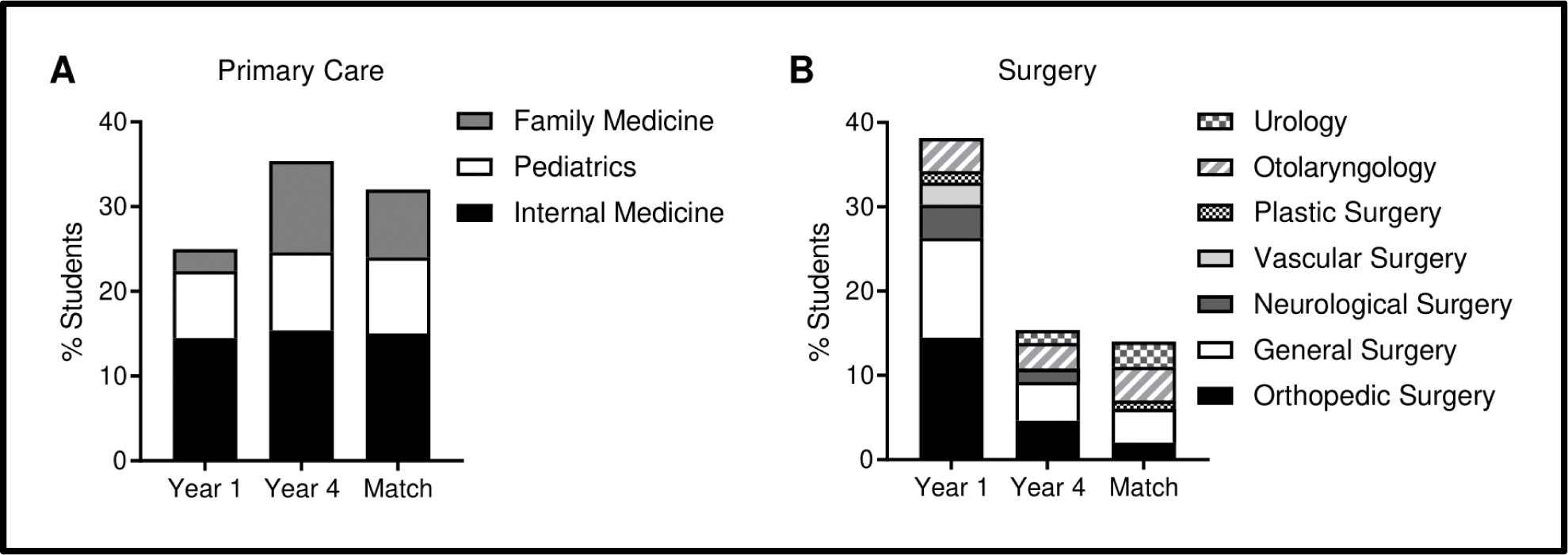
Percentage of students selecting primary care versus surgical residencies. **A)** Percentage of students selecting primary care specialities in Year 1, Year 4, and the Match outcome. **B)** Percentage of students selecting surgical specialities in Year 1, Year 4, and the Match outcome.

With the knowledge that considerable changes occurred between the proportion of students interested in primary care versus surgery in Year 1 compared to Year 4 and the Match, we next wanted to assess if and how our panel of motivational factors may have changed within primary care and surgery from Year 1 to Year 4. For primary care, interest in the field itself significantly increased as a factor motivating students’ choice of a career in a primary care specialty from Year 1 to Year 4, whereas prestige and financial incentives associated with primary care specialties decreased (**Figure 3A**). Specialty lifestyle, match confidence, and family/location factors did not significantly change. For surgery, interest in the field itself significantly increased from Year 1 to Year 4, while the influence of prestige decreased (**Figure 3B**). Factors pertaining to specialty lifestyle, financial incentive, match confidence, and family/location did not significantly change amongst those interested in surgery between Years 1 and 4.

**Figure 3.**
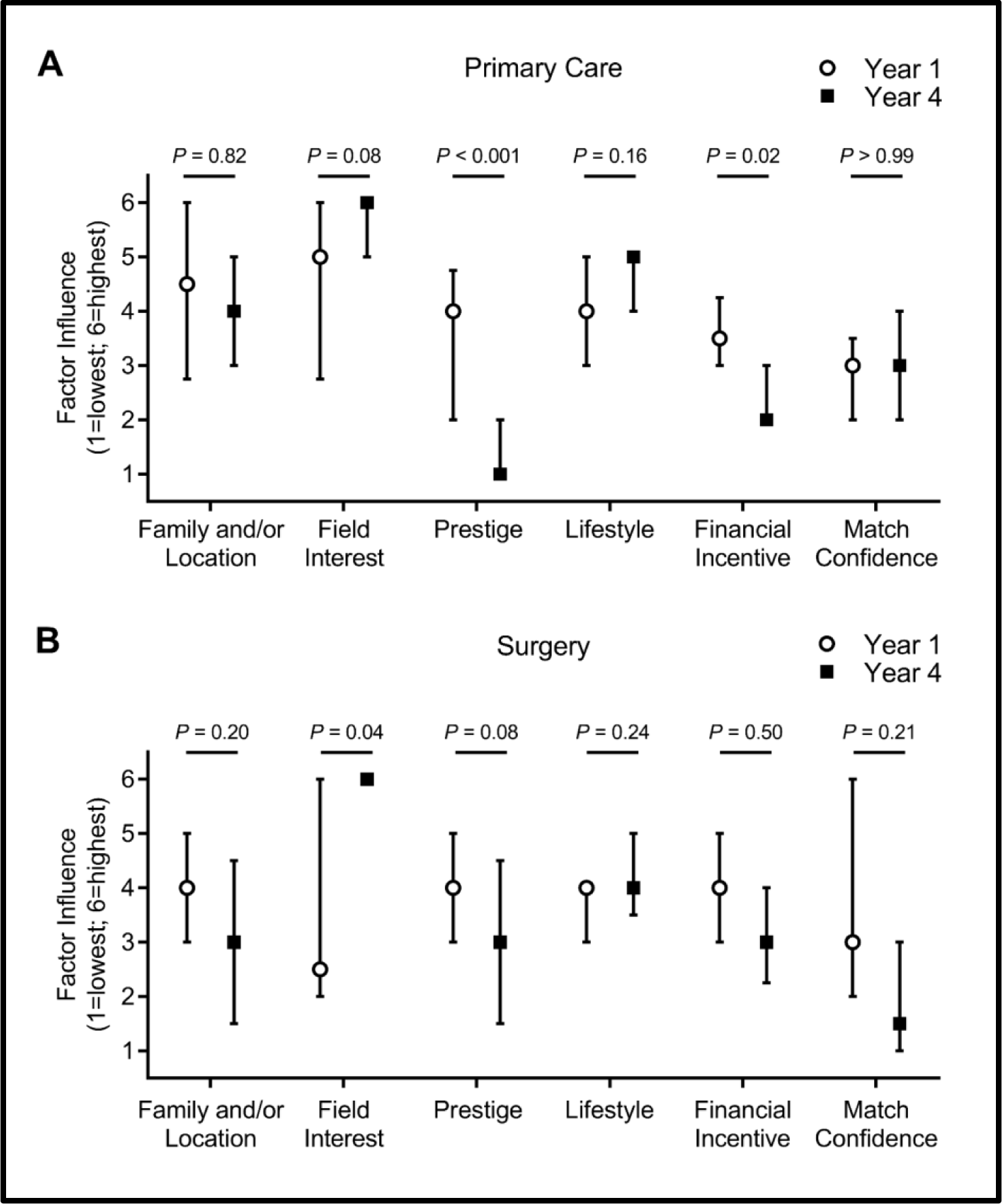
Factors influencing residency choice by year within primary care and surgery. Median and interquartile range for the six motivational factors influencing choice of residency specialties from least important (1) to most important (6), comparing **A)** the student cohort in Year 1 (white circle) versus Year 4 (black square) amongst those who chose primary care specialties, and **B)** the student cohort in Year 1 (white circle) versus Year 4 (black square) amongst those who chose surgical specialties. *P*-values were calculated using Mann-Whitney U tests.

Additional analyses were performed to assess if and how these motivational factors differ between primary care and surgery within Year 1 and Year 4. For Year 1, none of the surveyed factors demonstrated a significant difference between students who were interested in primary care versus surgery (**Figure 4A**); however, significant changes were observed in Year 4 (**Figure 4B**). The influence of family and/or location factors was significantly lower in students who had chosen a surgical specialty in Year 4 compared to primary care, as was the importance of confidently matching into the specialty of choice. Furthermore, specialty prestige and financial incentive were more influential amongst students who were pursuing a surgical specialty. Field interest and lifestyle were not significantly different between the two groups in Year 4.

**Figure 4.**
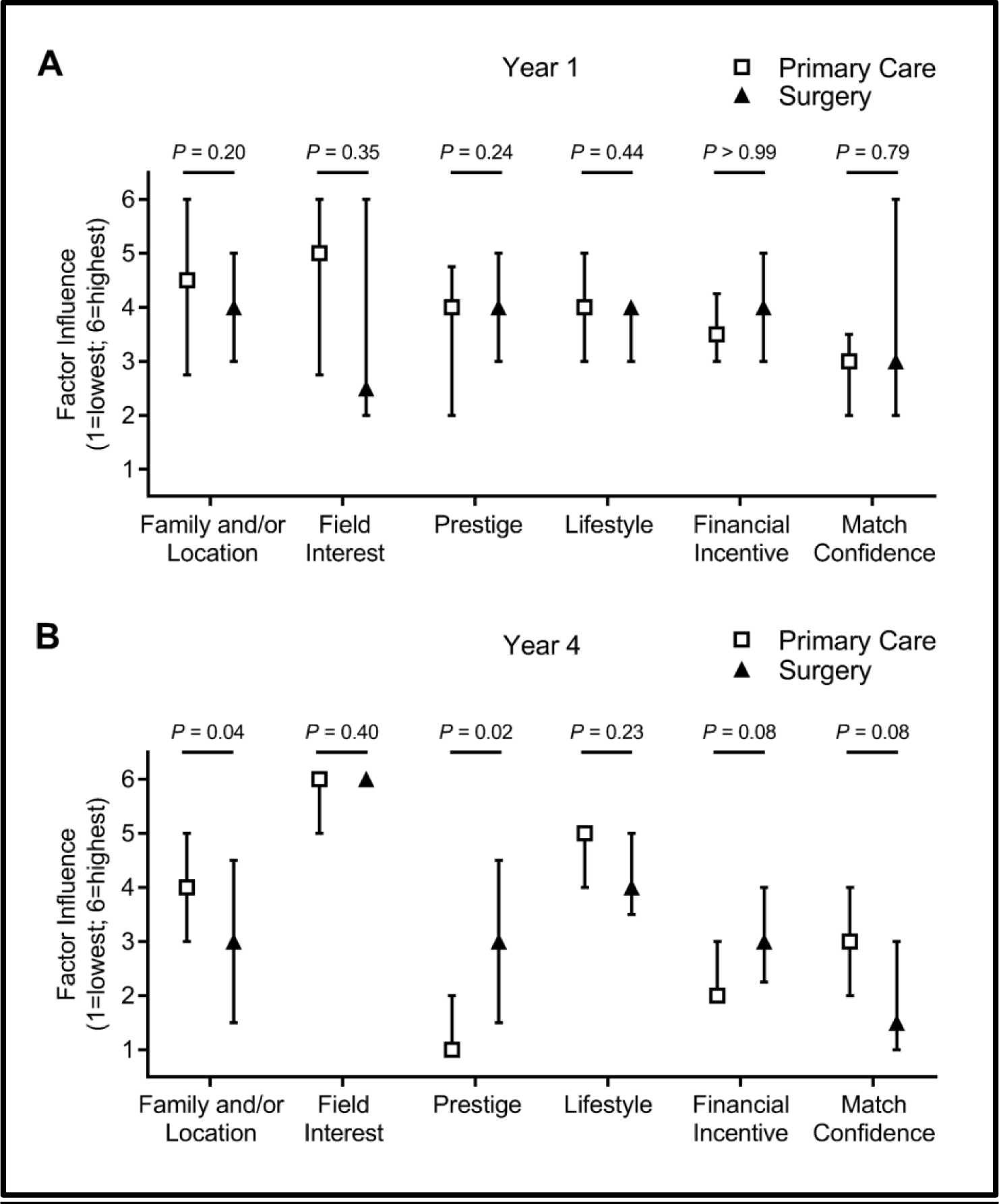
Factors influencing residency choice between primary care and surgery in Years 1 and 4. Median and interquartile range for the six motivational factors influencing choice of residency specialties from least important (1) to most important (6), comparing **A)** Year 1 medical students who chose primary care (white square) versus surgical (black triangle) specialties, and **B)** Year 4 medical students who chose primary care (white square) versus surgical (black triangle) specialties. *P*-values were calculated using Mann-Whitney U tests.

## Discussion

While students enter medical school with specific specialties they intend to pursue, which may be predictive of career choice in some studies (44,45), up to 70-80% of medical school graduates have been shown to ultimately pursue specialties different from those declared at matriculation, a figure that has persisted for many decades (26,46). Importantly, the worsening mismatch between physician supply and demand in key specialties hinges, in part, on the career decisions made by medical students. Thus, understanding these decisions can provide invaluable information that could be harnessed to shape training goals for both students and healthcare as a whole. As such, the overall aim of this study was to track how final residency choices differ from those at matriculation, and what factors may play a role in these differences.

In the cohort surveyed in this study, a considerable decrease was observed regarding the proportion of students choosing surgical specialties in their final year compared to matriculation. The top residency choices at matriculation were internal medicine (14.5%), emergency medicine (14.5%), orthopedic surgery (14.5%), general surgery (11.8%), pediatrics (7.9%), and OB/GYN (7.9%). In contrast, the top choices for this cohort in their final year were internal medicine (15.4%), emergency medicine (12.3%), family medicine (10.8%), pediatrics (9.2%), OB/GYN (9.2%), and radiology (9.2%). Notably, 25.0% of students at matriculation were interested in primary care, while 38.2% were interested in surgical specialties. In the final year, these results shifted to 35.4% and 15.4%, respectively. The specialty choices that exhibited the greatest absolute change were orthopedic surgery (−9.9%), family medicine (+8.1%), radiology (+7.9%), general surgery (−7.2%), and anesthesiology (+6.2%). The final year survey results were similar to the final NRMP Match results for this cohort.

These results are in accord with previous studies, which have demonstrated that ∼30% of medical graduates now pursue primary care, a globally applicable percentage that has been in decline across multiple countries (22,28,47,48). Similarly, a decline in graduating medical students choosing surgical specialties has been observed over the years, such that ∼15% of medical graduates now pursue surgery (6,7,49,50), in line with the percentage observed in our study. The other ∼55% of graduates mostly pursue emergency medicine, OB/GYN, radiology, anesthesiology, and psychiatry, in varying proportions. Moreover, the declining entry rates of students into primary care and surgery are generally inconsistent with student preferences upon matriculation, as it has been previously reported that students choose residencies different than the ones they claim to be interested in upon matriculation (21–25,46). Within this context, our study further supports this by providing the most current investigation of these multi-decade patterns that continue to afflict physician supply.

Importantly, our study additionally aimed to examine the underlying motivational factors that influence residency choice, with particular focus on how these factors change with time and specialty choice. Compared to matriculation, influence of specialty interest and lifestyle were significantly more important when students made their final residency choice, whereas field prestige, financial incentive, and match confidence were significantly less important. Similar temporal trends existed when results were stratified by residency choice amongst primary care and surgical specialties. As the influence of certain factors differed with time and coincided with an increase and decrease in the proportion of students choosing primary care and surgery, respectively, we wanted to test if factor differences existed between these specialty categories over time, as a frequently proposed theory regarding primary care being a less attractive career choice is due to the salary inequality compared to other specialities (51,52), as well as the lower perceived prestige (22,53). At matriculation in our cohort, no significant differences were detected amongst the factor rankings between students interested in primary care versus surgery; however, in the final year, students choosing surgery ranked prestige and financial incentive significantly higher than primary care. In contrast, final year students interested in primary care rated match confidence and family/location factors higher than students in surgery. These results support another previously proposed theory that lifestyle, hours, and training commitment are a common reason that students may shy away from surgical specialties (53–56). Finally, both groups expressed a similarly high importance of interest in their chosen field as well.

Interestingly, we believe that these results provide preliminary evidence that students may exhibit different risk-reward profiles based on the type of residency specialty they choose to pursue. Specifically, students interested in surgery may risk successfully matching into these more competitive residencies for the reward of the salary and perceived prestige that accompany many surgical specialties, whereas students pursuing primary care place greater emphasis on family/location requirements that may also relate to match confidence. Vice versa, students interested in surgery may have more competitive application characteristics, giving them greater confidence and having less concern about matching. While these implications would need to be more closely examined amongst larger student cohorts in order to establish more definitive correlation, the associations observed in our study are not without precedent. It has previously been reported, albeit in separate studies, that students pursuing primary care are more motivated by medical lifestyle/work-life balance (23,27,30), ease of residency entry (23), and family status (24). In contrast, students pursuing surgery or other non-primary care fields were more motivated by economics (23,24,28,29), prestige (22,24), and having more competitive application characteristics (23,57). Our study adds to these previous findings by comprehensively assessing the relative influence of many of these factors, not only on the basis of specialty choice, but also, uniquely, in association with different training stages.

However, while students state having high interest in the fields they pursue and contemplate additional key factors supporting that pursuit, many residents and physicians regret or are unsatisfied with their career choice (1,58,59), undermining the health of a long-depleted workforce (14–16). One reason, amongst many, that may explain this is that students may not fully understand the specialties they choose and how they align with their personal/career values. Additionally, the formal, informal, and hidden curricula students are exposed to through their interpersonal, organizational, and cultural interactions in medical school can have significant impact on students’ career perceptions (60,61). As our study shows, newly matriculated students interested in primary care versus surgery exhibit no differences in their ranking of motivational factors, but many of these factors significantly deviate between the two career paths in the final year, as we have described. This demonstrates, in part, the impact of the medical curriculum. While exposure to the informal and hidden curricula will vary from student-to-student and school-to-school, making it difficult to exactly measure and control, interventions regarding the formal curriculum have been well-established. Particularly, outside of the typical required clinical clerkships, longitudinal and auxiliary experiences have been shown to foster student interest and impact residency choice (24,27,28,31–33). The long-term effects regarding career choice regret and satisfaction have yet to be explored, but these experiential curricular additions offer an opportunity for medical schools to provide more personalized scholastic exposures that cultivate student interests, which could be performed in conjunction with consideration of student values upon matriculation and throughout major training stages.

### Study Limitations

This study has important limitations to consider. First, analysis was performed on a single U.S. medical student cohort from the University of Connecticut, a public research university with longitudinal curriculum experiences that provide significant exposure to primary care, making external extrapolation challenging, though multi-institutional studies have shown consistent residency choice outcomes between schools (22,30). Second, cohort sizes at this institution are roughly ∼100 students per year, and when considering factors that can dampen response rate for a voluntary survey (student availability, willingness, survey completion, etc.), our sample size was limited, and particularly low for Years 2 and 3, hindering our ability to confidently assess these stages due to non-response bias. This small class size failed to demonstrate interest (at matriculation or in the Match) in child neurology, combined internal medicine and pediatrics, pathology, and physical medicine and rehabilitation, likely owing to the roughly ≤1% national Match outcomes for these specialties (62). While the Year 4 survey results correlated well with our cohort’s Match outcomes, our limited response rate also failed to identify the high proportion of students who matched into psychiatry, illustrating potential confounding from non-response bias. Finally, while inspired by previous studies, our questionnaire format and delivery method can be improved upon. The anonymous nature of our survey prevented us from tracking the same students throughout the study, limiting more refined longitudinal analysis. Methods for anonymity through electronic questionnaire distribution could address this and also expand the cohort scope and sample size in future studies. Additionally, the ordinal scale of our ranking system limits weighted analysis of the motivational factors examined in this study. Allowing respondents to attribute influence weight to these ranked factors, as well as expanding the specificity and range of factors surveyed in the questionnaire, such as assessing students’ perceptions of specialty-specific physician burnout and reasons behind factors such as match confidence, could provide more accurate and nuanced results.

## Conclusion

In summary, this study examined the longitudinal residency choices and motivational factors for a cohort of U.S. medical students, with the aim of generating insight that could aid the training of the next generation of physicians. We identified how residency choices change between the beginning and end of medical school, how the influence of certain factors change over this period, and stratify our results by specialty choice between primary care and surgery. Our study promotes awareness of student preferences, provides a blueprint for future studies to examine these factors on a larger scale, proposes a new theory based on risk-reward balance regarding residency choice, and may help guide medical school curricula in developing more student-tailored approaches to education and training. Eventually, we hope this work can help play a part in addressing the supply and dissatisfaction issues plaguing the physician workforce, which will ultimately improve healthcare outcomes for patients.

## Data Availability

The acquired dataset used and analyzed during the current study are not publicly available due to participant anonymity but are available from the corresponding author on reasonable request.

## Declarations

## Abbreviations

IRB: Institutional Review Board
UConn SOM: University of Connecticut School of Medicine
ERAS: Electronic Residency Application Service
NRMP: National Resident Matching Program
AAFP: American Academy of Family Physicians
ACS: American College of Surgeons
OB/GYN: Obstetrics and gynecology

## Ethics approval and consent to participate

This study was reviewed and approved by the Human Subjects Institutional Review Board (IRB) at UConn Health (IRB number 18-062-3) and all participates were consented for the study. All methods were carried out in accordance with the institutional guidelines and regulations. All the participants provided informed consent to participate in the study.

## Consent for publication

Not applicable.

## Author Contributions

F.A.L. and A.M.P.: Conceptualization, Data curation, Formal Analysis, Investigation, Methodology, Project administration, Resources, Validation, Visualization, Writing – original draft, and Writing – review & editing. A.E.P.: Conceptualization, Methodology, Project administration, Supervision, and Writing – review & editing.

## Acknowledgements

We would like to thank the UConn School of Medicine students who volunteered to participate in this study, professors Raymond J. Foley, D.O. (UConn) and Jason W. Ryan, M.D., M.P.H. (UConn) for helping the authors coordinate appropriate times to distribute the questionnaires used in this study, and professor James J. Grady, Dr.P.H. (UConn) for consultation regarding statistical analyses. The authors were supported, in part, by fellowship training grants from the American Heart Association (PRE35110005 to F.A.L. and PRE34381021 to A.M.P.) and institutional funds from the UConn School of Medicine, Graduate School, and M.D./Ph.D. Program.

